# Regional and whole-brain neurofunctional alterations during pain empathic processing of physical but not affective pain in migraine patients

**DOI:** 10.1101/2025.02.16.25321714

**Authors:** Dan Liu, Menghan Li, Heng Jiang, Yiqi Mi, Xiaolei Xu, Yali Zhou, Yaxian Hou, Yuquan Shen, Michael Maes, Xiao Xiao, Feng Zhou, Luca Giani, Keith M. Kendrick, Benjamin Becker, Stefania Ferraro

**Author notes:** Corresponding Author: Stefania Ferraro Sichuan Provincial Center for Mental Health, Sichuan Provincial People’s Hospital, University of Electronic Science and Technology of China, Chengdu, China and Ministry of Education Key Laboratory for Neuroinformation, School of Life Science and Technology, University of Electronic Science and Technology, Chengdu, China.

## Abstract

**Background:** Accumulating evidence suggests that migraine patients present abnormal brain responses to salient sensory and emotional stimuli. However, it is still unclear whether this is a generalized or domain-specific phenomenon. Employing a well-validated fMRI paradigm, we investigated pain empathic reactivity across two domains: observation of physical pain (noxious stimulation) and affective pain (facial expressions). On the basis of a generalized hyperexcitability/hyperreactivity in migraine, we hypothesized abnormal responses to both dimensions of pain empathy.

**Methods:** We collected fMRI and psychometric data from 21 migraine patients and matched controls. Univariate and multivariate neuroimaging analyses were utilized to examine domain-specific dysregulations in (a) neural reactivity in meta-analytically defined shared regions of pain-empathy processing, and (b) whole-brain neurofunctional signatures of physical and affective pain empathy (VPS, Zhou et al., 2020). Logistic regression models and machine learning-based classification were employed to determine differences between groups (migraine or control).

**Results:** Migraine patients exhibit increased neural responses during empathy for physical pain in the bilateral inferior frontal gyrus (slightly more pronounced on the right side), with alterations on the right significantly associated with the pain experienced during the attack. On the whole-brain level, the predictive accuracy of the VPS for physical pain empathy was shown to be significantly higher for patients as compared to controls, reaching 100% accuracy. Across analyses, we did not find evidence of altered empathy processing for affective pain.

**Conclusion:** Contrary to our hypothesis, our results indicate that migraine patients present a domain-specific increased brain responsivity, localized in the bilateral inferior frontal gyrus but also extending to subtle whole brain patterns, during empathy for physical pain stimuli, but not during empathy for affective pain. Based on the evidence that the neural pathways for empathy for physical pain and experimental pain robustly overlap, these results indicate a specific hyperresponsivity of the pain pathways, with the inferior frontal gyrus likely playing a regulatory role in modulating pain-related processes. Finally, the results underscore the translational application potential of neuroaffective multivariate signatures as neuromarkers for pathological dysregulations in affective and pain-related processes.

## 1. Introduction

Migraine, a primary headache disorder that affects approximately up to 15% of the world’s population (Steiner et al. 2020) has been identified by the Global Burden of Disease Survey (2019) as the second leading cause of disability-adjusted life years (DALYs - calculated as years of life lost and lived with disability) in both sexes and all ages (Vos et al. 2020) and the first among women under 50 years of age (Steiner et al. 2020). Despite this enormous personal, social, and economic burden, the pathophysiology of this disorder remains poorly understood. Current evidence supports the hypothesis of a genetic predisposition (Sutherland, Jenkins, and Griffiths 2024) associated with a brain characterized by hyperexcitability*/*hyperreactivity or by an abnormal switching between hypo- and hyperexcitable states (Goadsby et al. 2017). Convergent evidence suggests that the hypothalamic and brainstem nuclei play a central role in the onset of the migraine attack. As key regulators of both physiological and emotional homeostasis and allostasis, these nuclei could lower the threshold for the transmission of nociceptive signals within the trigeminovascular system (Noseda et al. 2014; Coppola et al. 2016).

The conceptualization of abnormal brain excitability has been supported by evidence indicating that migraine patients, both during the ictal and interictal phases, exhibit a generalized alteration of sensitivity to sensory stimuli (Goadsby et al. 2017; Schwedt, Zuniga, and Chong 2015) and emotional experiences, although it remains unclear whether there is a predilection for negative affective stimuli (M. Wang et al. 2017; Wilcox et al. 2016; Steppacher, Schindler, and Kissler 2016; Andreatta et al. 2012; Szabó et al. 2019).

Given the inherently aversive nature of pain, encompassing both sensory and emotional dimensions, it is not surprising that pain perception in migraine is also abnormal. Indeed, despite some inconsistencies (Pan et al. 2020), patients with migraine were shown to present abnormal pain thresholds to different types of nociceptive stimulation (Schwedt et al. 2011; Beese et al. 2015; Sand et al. 2008; De Tommaso et al. 2014; Palacios-Cena et al. 2016; Pan et al. 2020; Nahman-Averbuch et al. 2018). Also from a neural perspective, migraine patients exhibit heightened responses to nociceptive stimuli (Russo et al. 2012; Schwedt, Zuniga, and Chong 2015) with increased antinociceptive activity (Kökönyei et al. 2021). This increased sensitivity extends to the emotional and cognitive dimensions of pain as well: severe headache amplifies fear of pain and fosters avoidance behaviors (Norton and Asmundson 2004) and directly impacts negative emotional states, which in turn influence migraine-related disability (Kim et al. 2021).

Empathy for pain, also referred here to as vicarious pain, is a multifaceted social-affective experience and refers to the capacity to resonate with and share others’ pain. The experience of vicarious pain is elicited by observing another person in pain, including witnessing the direct infliction of a noxious stimulation or facial expressions of pain (Decety and Ickes 2011; Zhou et al. 2020; X. Xu et al. 2020). Alterations in behavioral and neural pain empathic reactivity have been observed across a growing number of disorders characterized by altered brain functioning, including chronic pain and neuropsychiatric disorders (Vachon-Presseau et al. 2013; He et al. 2024).

In patients with pain-related disorders, the abnormalities not only affect the level of pain perception (Amiri et al. 2021) but also the level of empathy for pain, as shown in fibromyalgia (de Tommaso et al. 2019) and primary dysmenorrhea (Mu et al. 2021), thus suggesting that dysregulation in the experience of painful conditions and the capacity to respond empathically to others’ pain are linked.

In this regard, empathy for pain is hypothesized to elicit an internal representation that engages parts of the neural pathways also implicated in pain processing (Singer et al. 2004; Lamm, Decety, and Singer 2011; Timmers et al. 2018). A recent meta-analysis (Fallon, Roberts, and Stancak 2020) further corroborates this view, identifying the anterior insula cortex (aIC), anterior midcingulate cortex (aMCC), inferior frontal gyrus (IFG), and somatosensory areas as shared hubs of these two related processes. The identified pathways, in particular the aIC and to a certain extent the aMCC, overlap with regions identified as robust functional biomarkers for pain hyper-reactivity in chronic pain patients (Ferraro et al. 2022). More recent approaches have furthermore combined fMRI with machine learning-based predictive modeling for pain-related and affective processes, including pain (Neural Pain Signature - Wager et al. 2013) and vicarious pain (Vicarious Pain Signature or VPS, Zhou et al. 2020). Progress in the development of these neuroaffective decoders (or neuromarkers) has demonstrated that affective and pain-related mental processes are represented in and can be accurately quantified based on distributed neural representations (see e.g., Gan et al. 2024; Liu et al. 2024; D. H. Lee, Lee, and Woo 2025; Čeko et al. 2022). The VPS, an fMRI brain pattern of empathy developed by Zhou et al. 2020 (also employed in Corradi-Dell’Acqua et al. 2023), demonstrated not only high sensitivity to vicarious pain but also, crucially, to acute thermal pain. Similar conclusions were reached by another recent study (Li et al. 2024) showing that a predictive pattern developed to detect physical pain (Neural Pain Signature - Wager et al. 2013) was also sensitive to vicarious pain.

The neural bases of empathy for pain can be examined across two dimensions: the physical and the affective. Physical pain empathy triggered by the observation of noxious stimulation of body limbs is thought to be primarily associated with the sensory components of the observed pain (hereafter, physical pain empathy). Differently, affective pain empathy elicited by facial expressions of pain, salient emotional-communicative cues (Schirmer and Adolphs 2017), and requiring the observer to interpret the expresser’s pain experience (Hadjistavropoulos et al. 2011) is thought to primarily reflect the affective dimension of the observed pain (hereafter, affective pain empathy) (Li et al. 2024; Zhou et al. 2020; Xu et al. 2020). Consistent with this hypothesis, the predictive model (VPS) developed by Zhou et al. (2020) demonstrated partially dissociable neural representations for physical pain empathy and affective pain empathy (see also Xu et al. 2020 for a convergent approach). Moreover, coherently with the above subdivision, their study showed that the predictive model specifically developed for physical pain empathy was a more accurate predictor of acute thermal pain compared to the model designed to detect affective pain empathy.

The relevance for neuropsychiatric disorders of the different empathic domains of pain is further underlined by previous studies reporting associations between disorder- and neural empathy-specific domains of pain and emotional dysregulation, e.g. in response to facial expressions of pain in depression (but not in generalized anxiety, Xu et al. 2020) or opposite relationships with alexithymia (Li et al. 2019).

Within this robust theoretical and emerging empirical framework, we examined the neural bases of physical and affective pain empathy in a cohort of migraine patients compared to a control group using a well-validated fMRI paradigm (J. Li et al. 2019; X. Xu et al. 2020) with visually presented stimuli depicting both physical pain (i.e., noxious stimulation of body limbs) and affective pain (i.e., facial expressions of pain). Our primary objective was to determine whether individuals with migraine are characterized by a global abnormality in the processes of empathy for pain, as hypothesized on the basis of altered brain responsiveness to aversive stimuli, or whether they are instead characterized by a dissociation between the physical and affective dimensions of empathy for pain, thus indicating a privileged alteration of a specific dimension of this process. Given previous findings of altered sensory and emotional responses to pain and to negative affective stimuli, we hypothesized abnormal responses to both dimensions of pain empathy. Furthermore, based on accumulating evidence for the distributed representation of mental processes, including pain and pain empathy, we applied the VPS signature to determine the application of neurofunctional signatures to detect disorder-related alteration in the processes under examination.

## 2. Methods

### 2.1. Participants

Twenty-two headache patients were recruited through online communication channels and by posters displayed in hospital facilities and compared with 23 matched control subjects.

As an initial screening process, candidates answered a set of preliminary questions developed with a neurologist (co-author, L.G.) experienced in headache disorders (1. Are you between 18 and 65 years old? 2. How long have you been suffering from headaches? 3. How many attacks do you have per year?). Based on their responses, participants were included in the initial dataset according to the following inclusion criteria: 18-65 years old, a history of headache of at least 1 year, and a minimum of 1 headache attack per month. Eligible participants were invited to enter the Critical Care Clinical Decision Support System (CDSS 2.0) (Han et al. 2023) and provide the required information. This CDSS 2.0 is able to make a diagnosis of the headache type based on clinical information obtained through a human-computer conversation. This system has a very high level of sensitivity and specificity compared with the gold standard (2 neurologists experienced in headache disorders evaluating a combination of the CDSS 2.0 self-generated report and outpatient medical records) for migraine without aura (sensitivity *=* 0.76, specificity *=* 0.99) and migraine with aura (sensitivity *=* 0.96, specificity *=* 0.97) in the general population (Han et al. 2023). Among the subjects who filled out the CDSS 2.0, we consecutively selected all patients (n*=*22) diagnosed with migraine (with and without aura) or probable migraine, which is believed to have pathophysiological mechanisms similar to those of definitive migraine (Ashina et al. 2021).

At this stage, the inclusion criteria of migraine patients were stable headache characteristics for at least one year before study entry; while exclusion criteria were as follows: concomitant diagnosis of other primary or secondary headaches, cardiovascular disease, diabetes mellitus, hypertension, history of psychiatric and other neurological conditions, contraindications to MRI, the onset of a headache attack in the 24 hours before the MRI session, and use of medications in the last 24 hours. During headache attacks, most patients consumed NSAIDs (almost 70%), and a minority Acetaminophen, while the remaining ones did not use any pharmacological treatment (see Table 1). No patient was taking prophylactic therapy. Twenty-three control (CTRL) subjects were recruited through social media advertisements. After discarding 2 control subjects and 1 migraine patient because of head motions greater than 2 mm during MRI acquisitions, clinical and MRI data from 21 migraine (MIG) patients (6 males; mean age*=* 29±7.8 years) and 21 CTRL subjects (8 males, mean age*=* 29± 8.7 years) were analyzed. The study was approved by the local ethics committee, and written informed consent was obtained from each participant.

**Table 1.**
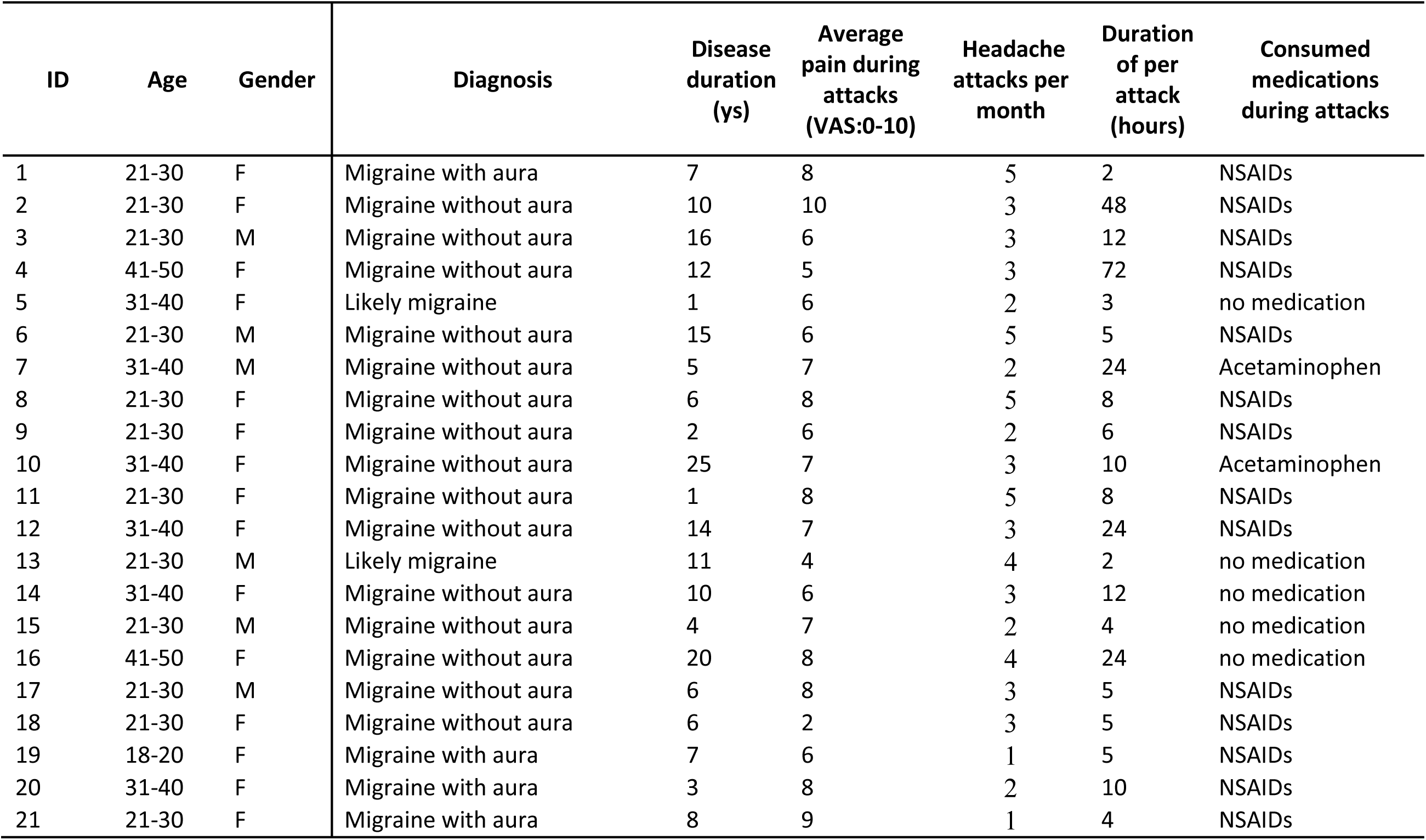
Demographic and clinical information for each migraine participant (final sample). Abbreviations: ys, years; VAS, visual analogue scale (0 no pain, 10 maximum imaginable pain); NSAIDs, non-steroidal inflammatory drugs.

| ID | Age | Gender | Diagnosis | Disease duration (ys) | Average pain during attacks (VAS:0-10) | Headache attacks per month | Duration of per attack (hours) | Consumed medications during attacks |
| --- | --- | --- | --- | --- | --- | --- | --- | --- |
| 1 | 21-30 | F | Migraine with aura | 7 | 8 | 5 | 2 | NSAIDs |
| 2 | 21-30 | F | Migraine without aura | 10 | 10 | 3 | 48 | NSAIDs |
| 3 | 21-30 | M | Migraine without aura | 16 | 6 | 3 | 12 | NSAIDs |
| 4 | 41-50 | F | Migraine without aura | 12 | 5 | 3 | 72 | NSAIDs |
| 5 | 31-40 | F | Likely migraine | 1 | 6 | 2 | 3 | no medication |
| 6 | 21-30 | M | Migraine without aura | 15 | 6 | 5 | 5 | NSAIDs |
| 7 | 31-40 | M | Migraine without aura | 5 | 7 | 2 | 24 | Acetaminophen |
| 8 | 21-30 | F | Migraine without aura | 6 | 8 | 5 | 8 | NSAIDs |
| 9 | 21-30 | F | Migraine without aura | 2 | 6 | 2 | 6 | NSAIDs |
| 10 | 31-40 | F | Migraine without aura | 25 | 7 | 3 | 10 | Acetaminophen |
| 11 | 21-30 | F | Migraine without aura | 1 | 8 | 5 | 8 | NSAIDs |
| 12 | 31-40 | F | Migraine without aura | 14 | 7 | 3 | 24 | NSAIDs |
| 13 | 21-30 | M | Likely migraine | 11 | 4 | 4 | 2 | no medication |
| 14 | 31-40 | F | Migraine without aura | 10 | 6 | 3 | 12 | no medication |
| 15 | 21-30 | M | Migraine without aura | 4 | 7 | 2 | 4 | no medication |
| 16 | 41-50 | F | Migraine without aura | 20 | 8 | 4 | 24 | no medication |
| 17 | 21-30 | M | Migraine without aura | 6 | 8 | 3 | 5 | NSAIDs |
| 18 | 21-30 | F | Migraine without aura | 6 | 2 | 3 | 5 | NSAIDs |
| 19 | 18-20 | F | Migraine with aura | 7 | 6 | 1 | 5 | NSAIDs |
| 20 | 31-40 | F | Migraine with aura | 3 | 8 | 2 | 10 | NSAIDs |
| 21 | 21-30 | F | Migraine with aura | 8 | 9 | 1 | 4 | NSAIDs |

### 2.2. Demographic and Clinical Data and psychometric assessments

All the participants were evaluated with the following psychometric scales: Beck Depression Inventory (BDI-II) (Beck, Steer, and Brown 1996), State-Trait Anxiety Inventory (i.e., STAI-S and STAI-T) (Spielberger 1970), Pain Catastrophizing Scale (PCS) (Sullivan, Bishop, and Pivik 1995), Emotion Regulation Questionnaire (ERQ) (Wang et al. 2007), Basic Empathy Scale (BES) (Jolliffe and Farrington 2006), and Toronto Alexithymia Scale (TAS) (Bagby, Parker, and Taylor 1994).

Clinical variables for MIG patients (number of headaches per month, number of years with migraine, the average migraine pain measured on a visual analogue scale with a range of 0-10, and consumed medications to control migraine pain) were also collected.

Depending on the normality of data distribution (assessed using the Shapiro-Wilk test), parametric tests (independent samples t-test) or non-parametric tests (Mann-Whitney U test, Chi-square test) were used to assess group differences in age, sex, and psychometric assessments. Given that previous studies have demonstrated that MIG patients exhibit higher BDI (Chu et al. 2018) and PCS (Kim et al. 2021; Kocakaya et al. 2023) scores compared to CTRL subjects and that pain-related disorders such as fibromyalgia and osteoarthritis can present a higher level of empathy and pain empathy (Fallon et al. 2015; Zhao et al. 2022), one-tailed t-tests were applied for these variables. A significance level of *p* <0.05 was set for all analyses. All the statistics in the manuscript were computed using JASP (v0.19.0.0) unless otherwise specified. The between-groups significant variables were used in subsequent analyses.

### 2.3. Experimental fMRI procedure

The present study used a validated fMRI paradigm to elicit the neural processes of empathy for pain (J. Li et al. 2019; X. Xu et al. 2020; Zhou et al. 2020). The entire experimental paradigm included a total of 64 images (each presented for 3 s) divided into 16 blocks (4 images per block) for 4 different conditions: in 2 conditions, participants were presented with images of noxious stimulation of body limbs inducing empathy for physical pain (PP) (e.g., a scene of cutting a finger with scissors) and the respective control condition (PPc; e.g., a scene of cutting a piece of paper with scissors), while in the other two conditions participants were presented with images showing facial expressions of pain, inducing empathy for affective pain (AP), and the respective control condition (APc; e.g., neutral facial expressions). The stimuli inducing empathy for affective pain were images of facial expressions of adult Chinese individuals and were gender balanced. In the same block, the stimuli were interspersed with a fixation cross lasting 1s, while the interval between blocks averaged 10 s (range: 8-12 s). As in previous work, the present study required participants to attentively watch the stimuli (Xu et al. 2020).

### 2.4. MRI Data Acquisition

MRI data were acquired with a 3.0 T Philips Ingenia system using a 32-channel coil. High-resolution structural 3D T1-weighted (T1w) image (TR *=* 8.19 ms, TE *=* 3.78 ms, FOV *=* 240 × 240 mm, flip angle *=* 8°, voxel size *=* 1 × 1 × 1 mm) and T2* 2D multi-slice echo-planar imaging (EPI) (TR *=* 2000 ms, TE *=* 30 ms, FOV*=* 240 × 240 mm, flip angle *=* 80°, voxel size 3 × 3 × 3.2 mm, 34 axial slices, 3 dummy volumes, 220 volumes collected) were acquired in each participant.

#### 2.4.1. Regions of interest (ROIs) analyses

To study group differences in specific brain regions involved in empathy processing for pain, we built regions of interest (ROIs) centered in the MNI coordinates of brain areas identified as key hubs of pain empathy processing in a recent neuroimaging meta-analysis (Fallon, Roberts, and Stancak 2020). To this aim, we used MarsBar 0.45 (Brett et al. 2002) to construct 10-mm spherical ROIs centered in the MNI coordinates of the following regions (as reported in the meta-analysis): the aMCC, the bilateral aIC, IFG, supramarginal gyrus (SMG), and the right superior parietal lobule (SPL) (see Figure 1 and Table 2). Notably, we used the MNI coordinates reported in the selected meta-analysis but we used the nomenclature of the regions as reported in the Automated Anatomical Labeling (AAL3) atlas (Rolls et al. 2020). These ROIs were used in the subsequent analyses as specified.

**Figure 1:**
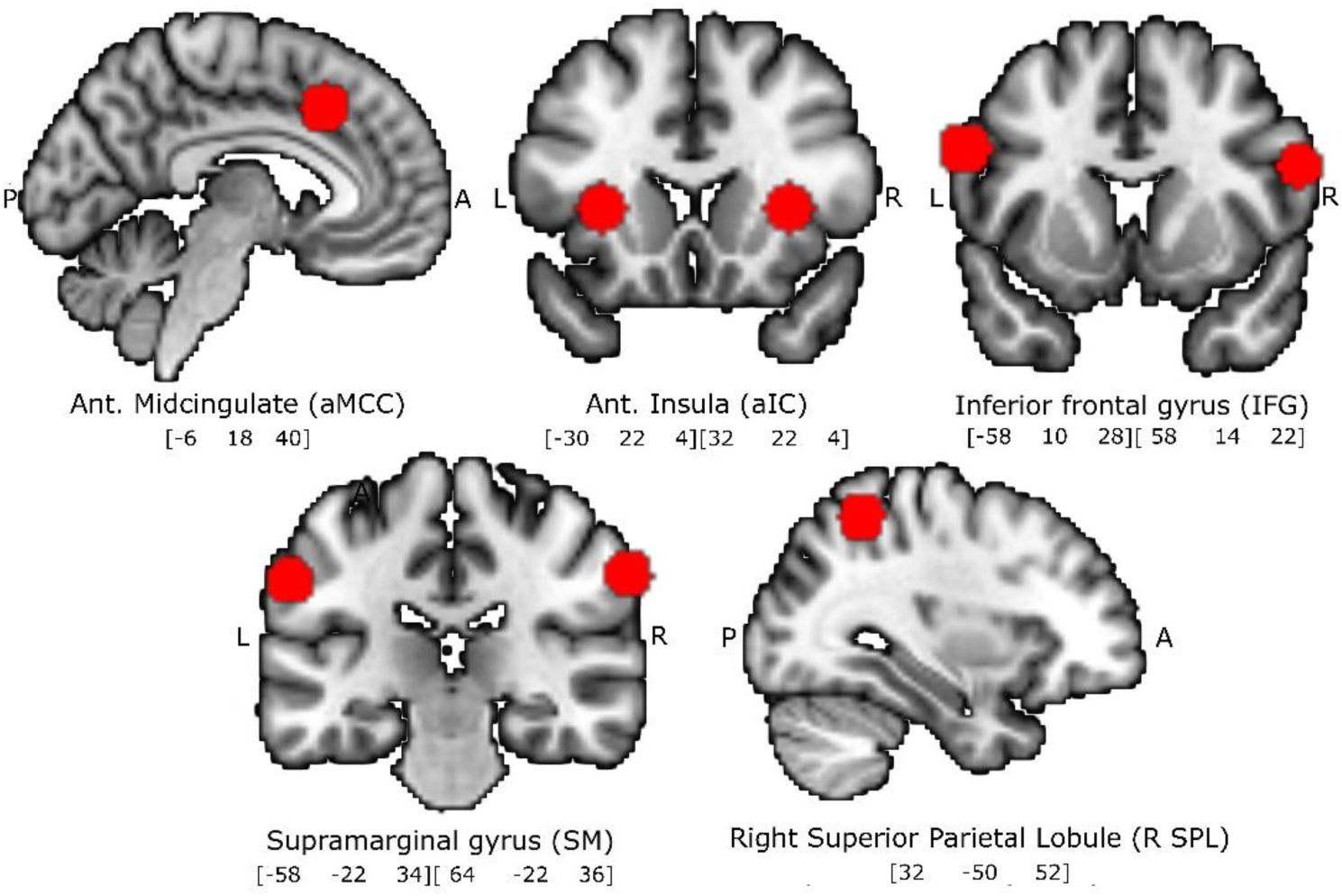
Representation of the 10mm spherical regions of interest (ROIs) employed for the extraction of the beta values obtained from the first-level fMRI analyses. ROIs were built using MarsBar 0.45 (Brett et al. 2002), and they were centered in the MNI coordinates identified from an independent neuroimaging meta-analysis on empathy for pain studies (Fallon et al., 2020).

**Table 2.** Regions of interest (ROIs): center coordinates. ROIs were built as 10mm spheres centered in the MNI coordinates, as reported in the meta-analysis of Fallon et al. (2020). Note that the attribution of the location of the coordinates to the respective brain region was done according to the Automated Anatomical Label atlas (AAL3) (Rolls et al., 2020).

| Brain region | Coordinates (MNI space) |  |  |
| --- | --- | --- | --- |
|  | x | y | z |
| Anterior midcingulate cortex (aMCC) | -6 | 18 | 40 |
| Left insula - anterior section (L aIC) | -30 | 22 | 4 |
| Left inferior frontal gyrus (L IFG) | -58 | 10 | 28 |
| Left supramarginal gyrus (L SM) | -58 | -22 | 34 |
| Right insula - anterior section (R aIC) | 32 | 22 | 4 |
| Right supramarginal gyrus (R SM) | 64 | -22 | 36 |
| Right inferior frontal gyrus (R IFG) | 58 | 14 | 22 |
| Right superior parietal lobule (R SPL) | 32 | -50 | 52 |

### 2.5. fMRI data preprocessing and statistical analyses

All functional and structural image data were preprocessed using SPM12 (Wellcome Department of Imaging Neuroscience, University College, London, UK; http://www.fil.ion.ucl.ac.uk/spm) running on MATLAB R2022a (The Mathworks, Inc.). For the functional images, after discarding the first 6 volumes, fMRI data were slice-timing corrected and spatially realigned to the first volume. The T1w image was segmented and spatially normalized to the Montreal Neurological Institute (MNI) space, and the produced parameters were then applied to the T1-w realigned fMRI data for normalization. All functional volumes were then spatially smoothed with an 8 mm full-width half-maximum (FWHM) Gaussian kernel. For each participant, the preprocessed functional data were entered into a first-level analysis in the framework of the general linear model (GLM). We modeled 4 regressors: empathy for affective pain (AP) and the respective control (APc), empathy for physical pain (PP), and the respective control (PPc). Motion parameters were used as predictors of no interest. Contrast images for the 4 regressors (AP, APc, PP, and PPc) were generated for use in the second-level whole-brain analysis. Additionally, the contrast images for AP>APc and PP>PPc were created to extract the corresponding beta values from the selected ROIs.

#### 2.5.1. Whole brain analysis

We employed a full factorial design with ‘condition’ (i.e., AP, APc, PP, PPc) as the within-subject factor and ‘group’ (MIG, CTRL) as the between-subject factor and we computed the main effect of ‘group’ and of the ‘condition’ and the interactions between ‘condition’ and ‘group’. Results were deemed significant for p-FDR<0.05.

#### 2.5.2. ROIs beta values analyses

Group differences were tested employing the average beta values extracted from each participant (for each ROI and contrast image) using the Matlab getbetas.m script (https://github.com/scanUCLA/spm12-getBetas) and applying the relevant statistics. For each ROI, a binary logistic regression model was used to test the statistical association between the extracted beta values from the two contrasts of interest (i.e., AP > APc and PP > PPc) and the diagnostic groups (MIG and CTRL).

For each ROI, a binary logistic regression model was used to test the statistical association between the diagnostic groups (MIG and CTRL, with the dependent variable coded as 1 for MIG and 0 for CTRL, the reference group) and the extracted beta values from the two contrasts of interest (AP > APc and PP > PPc), which were included as predictors in the model.

To rule out the possibility that the significant results of the logistic regression models were influenced by demographic and psychometric variables, we performed, for each significant ROI-i.e. the ROIs for which the beta values of at least one contrast were significant-2 linear regression analyses (one for each contrast) using the beta values as the dependent variable and the demographic data and/or psychometric scores that significantly differentiated MIGs from CTRLs as predictors. As the last step, in the patient group, we correlated the beta values of the contrast of interest (e.g., PP>PPc and AP>APc) with the disease duration and the average pain during migraine attacks. As for the linear regression models, we performed this computation only for the ROIs in which beta values were significant in the logistic regression analyses. Results were deemed significant for p<0.05 for all the conducted analyses.

#### 2.6. Vicarious Pain Signature (VPS): predicting affective (AP) and physical pain (PP) empathy in MIG and CTRL group

To strengthen the specificity of the present findings, we exploited existing multivariate brain patterns for pain empathy, applying the vicarious pain signature (VPS, developed by Zhou et al. 2020) to our fMRI dataset in the two diagnostic groups (i.e., MIG and CTRL) separately (for a similar approach, see also R. Zhang et al. 2023). More specifically, we used the VPS maps of facial expression vicarious pain (or FE-VPS) and of noxious stimulation vicarious pain (or NS-VPS) to test whether these multivariate brain patterns were able to predict the vicarious pain stimulation in our dataset. Specifically, the FE-VPS was used to predict the AP (i.e., affective empathy) condition relative to its control (i.e., AP vs. APC), and the NS-VPS to predict the PP (i.e., physical empathy) condition relative to its control (PP vs. PPc).

To evaluate these relationships, we employed the CANlab Core Tools (https://github.com/canlab/). First, we computed the dot-product between FE-VPS and the vectorized contrast images, obtained from our first-level fMRI analyses, for affective pain empathy and its respective control (AP and APc con images). The same approach was applied between the NS-VPS and the vectorized contrast images for physical pain empathy and its respective control (PP and PPc con images). This yielded a pattern expression value for each subject and condition, reflecting the similarity between the subject’s brain activation during the different contrast and the respective VPS predictive pattern. The pattern expression values were then used to assess the classifier’s discriminative performance between the target conditions (AP vs. APc, and PP vs. PPc). A Receiver Operating Characteristic (ROC) analysis was conducted separately for affective pain and physical pain brain activity, with the VPS serving as predictors and the binary condition labels as outcomes. The area under the curve (AUC) was calculated as a measure of the classifier’s accuracy in distinguishing the conditions (AP vs. APc, PP vs. PPc), along with significance testing (p-value). This approach allowed us to quantify how well the VPS predictive patterns captured the neural signatures associated with affective and physical pain empathy across subjects in the group of MIG patients and in the group of CTRL subjects.

Next, to determine whether the classification accuracy of FE-VPS and NS-VPS observed in the MIG group was significantly different from the accuracy expected under the null hypothesis, we generated a null distribution of classification accuracy values through 10,000 permutations. In each permutation, subjects from the MIG or CTRL groups were randomly reassigned, and the ROC analysis was recalculated along with the corresponding prediction accuracy.

Moreover, to improve comparability between subjects and minimize the impact of potential outliers, we rescaled all mean beta images to the same scale using Z-score normalization. The observed accuracy for the MIG group was then compared to the null distribution to compute the empirical p-values.

## 3. Results

### 3.1. Demographic and Clinical Data, and Psychometric Assessments

Demographic and clinical variables and relative statistics are shown in Tables 1 and 3. All the MIG patients in our sample were characterized by having 1 to 5 attacks per month. No differences were detected between groups for age and sex composition. MIG patients differed significantly from the CTRL group in BDI (one-tailed: U = 142, p = 0.024), PCS (one-tailed: U = 80.5, p < 0.001), and the cognitive empathy subscale of BES (U *=* -147.5, *p =* 0.049) (see Figure 2). No differences between groups were detected for the remaining variables. Only BDI, PCS, and BES scores were used in the subsequent analyses where explicitly stated.

**Figure 2:**
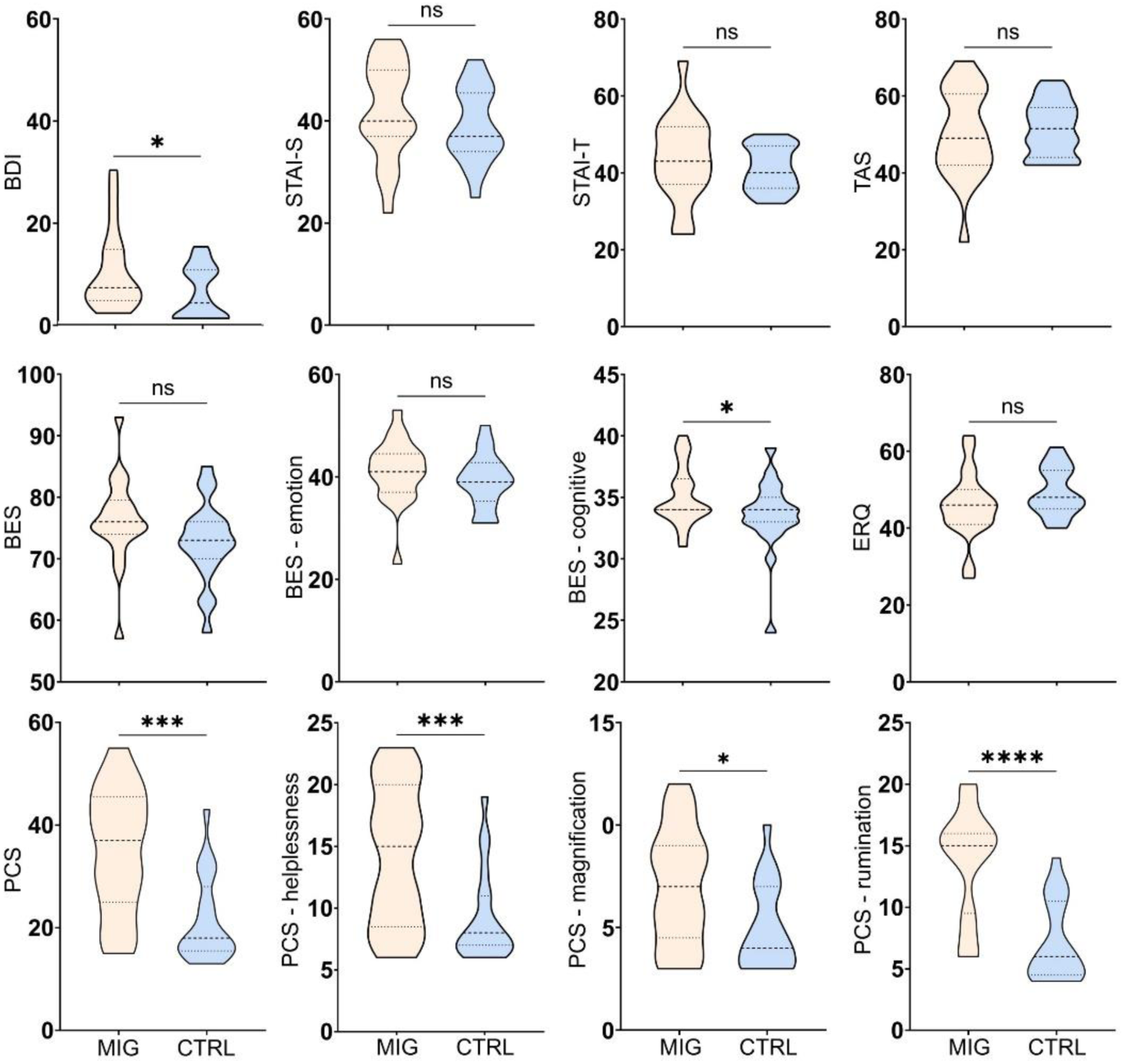
Violin plots of the psychometric assessments. Significant p-value: *p<0.05, ** p< 0.001, ****: p<0.0001. Abbreviations: ns, non-significative; BDI, Beck Depression Inventory; BAS, Basic Empathy Scale; EQR, Emotion Regulation Questionnaire; PCS, Pain Catastrophizing Scale; STAI-S, State Anxiety Inventory; STAI-T, Trait Anxiety Inventory; TAS, Toronto Alexithymia Scale.

**Table 3.** Between-group comparisons of demographic and clinical variables. Mann-Whitney test was applied when there was a violation of the normality of data distribution (assessed using the Shapiro-Wilk test). For the Student t-test, effect size was computed as Cohen’s d; for the Mann-Whitney test as the rank biserial correlation. *one-tail test. Abbreviation: MIG, migraine patients; CTRL: control subjects; VAS, visual analogue scale; BDI, Beck Depression Inventory; BAS, Basic Empathy Scale; EQR, Emotion Regulation Questionnaire; PCS, Pain Catastrophizing Scale; STAI-S, State Anxiety Inventory; STAI-T, Trait Anxiety Inventory; TAS, Toronto Alexithymia Scale.

|  | MIG | CTRL | Statistic | Effect Size | P-value |
| --- | --- | --- | --- | --- | --- |
| Sex (M/F) | 6/15 | 8/13 | $\chi^2(1) = 0.43$ | 0.101 | 0.513 |
| Age (M±SD) | 28.8±7.82 | 28.6±8.66 | $t(40)=-0.07$ | 0.023 | 0.941 |
| Duration of migraine (years, M±SD) | 9±6.14 | - | - | - | - |
| Pain level during attacks - VAS: 0-10 (M±SD) | 6.8±1.72 | - | - | - | - |
| Duration of single episode (hours) | 13.9±16.8 | - | - | - | - |
| BDI score (M±SD) | 9.3±7.95 | 5.0±5.12 | $U=142$ | -0.356 | 0.024* |
| BES score (M±SD) | 76.2±6.93 | 72.8±6.56 | $t(39)=-1.65$ | -0.517 | 0.053* |
| BES - Cognitive empathy (M±SD) | 35.1±2.39 | 33.6±2.98 | $U=-147.5$ | -0.298 | 0.049* |
| BES - Emotion empathy (M±SD) | 41.1±6.23 | 39.2±5.26 | $t(39)=-1.08$ | -0.336 | 0.144* |
| ERQ score (M±SD) | 45.9±8.67 | 49.5±6.12 | $t(40)=-1.54$ | 0.476 | 0.131 |
| PCS score (M±SD) | 35.0±11.98 | 21.4±8.24 | $U=80.5$ | -0.696 | < 0.001* |
| PCS subscale - Rumination (M±SD) | 13.8±4.46 | 7.2±3.2 | $U=51.5$ | -0.825 | < 0.001* |
| PCS subscale - Magnification (M±SD) | 7.0±2.81 | 5.0±2.06 | $U=129.5$ | -0.442 | 0.011* |
| PCS subscale - Helplessness (M±SD) | 14.3±5.63 | 9.2±3.68 | $U=99$ | -0.542 | 0.001* |
| STAI -S score (M±SD) | 41.9±9.38 | 39.1±7.25 | $t(40)=1.09$ | -0.335 | 0.284 |
| STAI-T score (M±SD) | 43.8±11.08 | 41.1±5.73 | $t(40)=0.99$ | -0.308 | 0.325 |
| TAS score (M±SD) | 50.1±11.66 | 51.6±7.09 | $t(39)=-0.50$ | 0.155 | 0.623 |

### 3.2. fMRI data results

#### 3.2.1. Whole-brain analysis results

The group-level analyses revealed a widespread network of brain regions activated for the main effect of ‘condition’ (see Table 1 SM and Figure 1 SM). However, no significant results were observed for the main effect of ‘group’, and for the interactions between ‘condition’ and ‘group’. For illustrative purposes, we conducted an analysis for the contrast AP+PP separately for the two groups, which revealed significant modulation in a set of bilateral regions, including the inferior frontal gyrus (IFG), extending to the right anterior insula cortex (R aIC), and occipital areas, with marked lateralization to the left supramarginal gyrus activity (see Figure 3).

**Figure 3:**
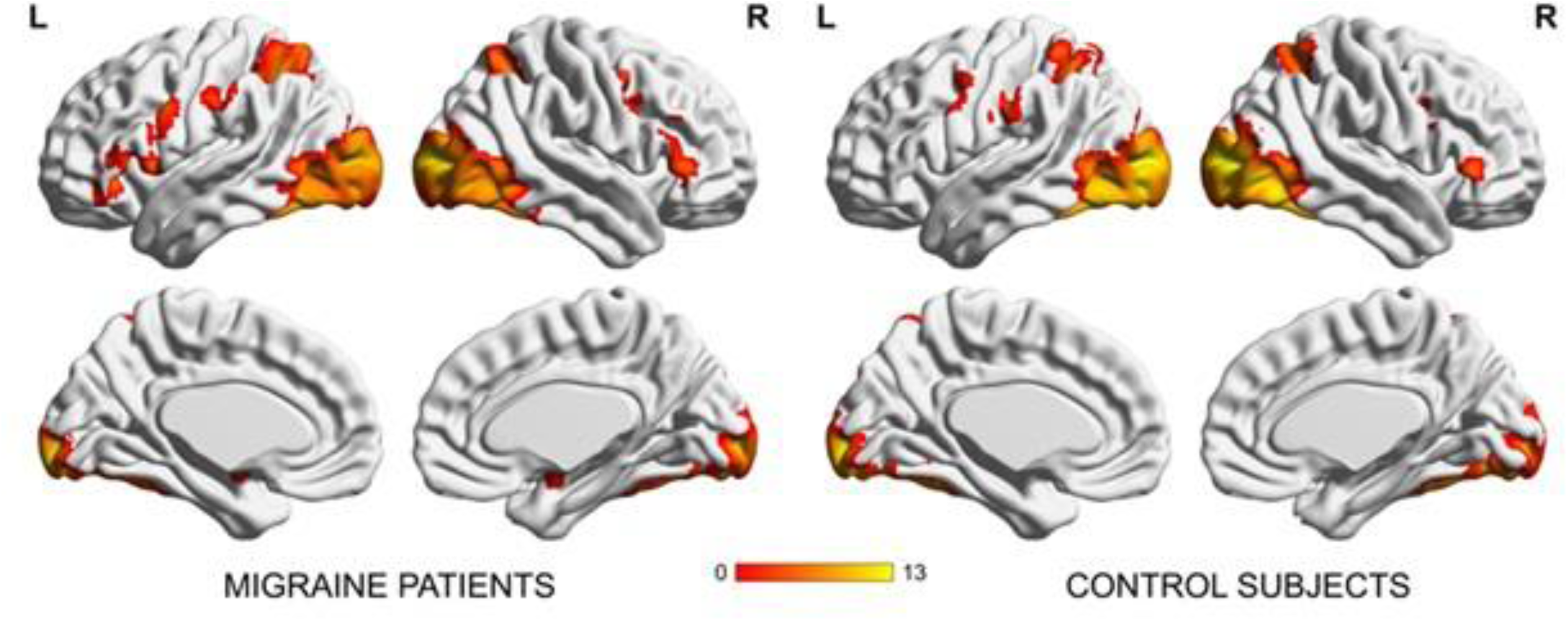
For illustrative purposes, we conducted a second-level analysis to identify the whole-brain fMRI activity during empathy for pain (contrast AP+PP) for each group (migraine and control). The color scale represents the T-values, with warmer colors (e.g., red and yellow) indicating higher T-values. Results were deemed significant for p-FDR<0.05.

#### 3.2.2. ROIs beta value results

Before applying logistic regression models, we computed the Variance Inflation Factor (VIF) among predictors and we found no significant collinearity (VIF for each predictor <2).

Logistic regression models (see Table 4) showed that the beta values of the contrasts of interest (i.e., AP > APc, PP > PPc) extracted from the R IFG [χ^2^(39) *=* 6.56, p *=* 0.038, Nagelkerke R² *=* 0.19] successfully discriminated between the diagnoses (i.e., MIG and CTRL). Similarly, the beta values extracted from the L IFG [χ^2^(39)*=* 5.97, p *=* 0.052, Nagelkerke R² *=* 0.177] were marginally significant. Among the two contrasts (see Table 5), only the PP > PPc significantly discriminated between the diagnoses in both 10mm spherical ROIs (R IFG: Wald test *=* 4.91, p *=* 0.027; L IFG: Wald test *=* 5.56, p *=* 0.033). No significant results were obtained for all the remaining ROIs.

**Table 4.** Summary of logistic regression models for each brain region of interest to estimate the probability of the diagnosis. The dependent variable was the diagnosis, coded as 1 for MIG (the group of interest) and 0 for CTRL (the reference group). Each model employed the beta values of the two contrasts for affective pain empathy and physical pain empathy (respectively, AP>APc, PP>PPc) extracted from the selected ROIs as predictors. * Significant p-value (p< 0.05). MIG, migraine patients; CTRL, control participants; aMCC, anterior midcingulate cortex; aIC, anterior insula cortex, SM, supramarginal gyrus; SPL, superior parietal lobule; IFG, inferior frontal gyrus; L, left; R, right; AUC, area under the curve; Sens, sensitivity; Spec, specificity; Prec, precision.

| MIG vs CTRL |  |  |  |  |  |  |  |  |
| --- | --- | --- | --- | --- | --- | --- | --- | --- |
| ROI | df | X<sup>2</sup> | p-value | Nagelkerke<br>R<sup>2</sup> | AUC | Sens | Spec | Prec |
| aMCC | 39 | 1.027 | 0.598 | 0.032 | 0.567 | 0.476 | 0.524 | 0.500 |
| L aIC | 39 | 1.988 | 0.370 | 0.062 | 0.612 | 0.571 | 0.619 | 0.600 |
| L SM | 39 | 2.208 | 0.332 | 0.068 | 0.608 | 0.524 | 0.619 | 0.579 |
| L IFG | 39 | 5.966 | 0.052 | 0.177 | 0.710 | 0.619 | 0.667 | 0.650 |
| R aIC | 39 | 1.792 | 0.408 | 0.056 | 0.649 | 0.762 | 0.429 | 0.571 |
| R IFG | 39 | 6.561 | 0.038* | 0.193 | 0.707 | 0.571 | 0.714 | 0.667 |
| R SPL | 39 | 3.425 | 0.180 | 0.104 | 0.667 | 0.714 | 0.619 | 0.652 |
| R SM | 39 | 2.346 | 0.309 | 0.072 | 0.540 | 0.429 | 0.524 | 0.474 |

**Table 5.** Coefficients of logistic regression models for each brain region of interest to estimate the probability of the diagnosis. The dependent variable was the diagnosis, coded as 1 for MIG (the group of interest) and 0 for CTRL (the reference group). * Statistically significant p-values (p<0.05). Abbreviations: MIG, migraine patients; CTRL, control participants; aMCC, anterior midcingulate cortex; aIC: anterior insula cortex; SM, supramarginal gyrus; SPL: superior parietal lobule, IFG, inferior frontal gyrus; L, left; R, right; OR, odds ratios; LB: lower boundary; UB: upper boundary; CI: confidence interval.

MIG vs CTRL
|  | Parameters | b | SE | OR | LB (95% CI) | UB (95% CI) | z | Wald Test | p-value |
| --- | --- | --- | --- | --- | --- | --- | --- | --- | --- |
| aMCC | (Intercept) | -0.054 | 0.327 | 0.947 | -0.696 | 0.588 | -0.165 | 0.027 | 0.869 |
|  | AP>APc | 0.883 | 1.286 | 2.417 | -1.639 | 3.404 | 0.686 | 0.471 | 0.493 |
|  | PP>PPc | 0.933 | 1.322 | 2.541 | -1.658 | 3.523 | 0.706 | 0.498 | 0.480 |
| L aIC | (Intercept) | -0.085 | 0.327 | 0.918 | -0.726 | 0.556 | -0.261 | 0.068 | 0.794 |
|  | AP>APc | 1.922 | 1.960 | 6.835 | -1.92 | 5.764 | 0.981 | 0.961 | 0.327 |
|  | PP>PPc | 2.53 | 2.545 | 12.549 | -2.458 | 7.517 | 0.994 | 0.988 | 0.320 |
| L SM | (Intercept) | -0.366 | 0.404 | 0.693 | -1.159 | 0.426 | -0.906 | 0.821 | 0.365 |
|  | AP>APc | 0.653 | 1.272 | 1.922 | -1.839 | 3.146 | 0.514 | 0.264 | 0.607 |
|  | PP>PPc | 2.377 | 1.689 | 10.773 | -0.932 | 5.687 | 1.408 | 1.982 | 0.159 |
| L IFG | (Intercept) | -0.335 | 0.360 | 0.715 | -1.041 | 0.370 | -0.931 | 0.868 | 0.352 |
|  | AP>APc | 0.970 | 1.060 | 2.639 | -1.107 | 3.048 | 0.915 | 0.838 | 0.360 |
|  | PP>PPc | 3.772 | 1.767 | 43.476 | 0.309 | 7.236 | 2.135 | 4.557 | 0.033* |
| R aIC | (Intercept) | -0.024 | 0.354 | 0.976 | -0.717 | 0.669 | -0.068 | 0.005 | 0.946 |
|  | AP>APc | 1.933 | 1.523 | 6.913 | -1.053 | 4.919 | 1.269 | 1.611 | 0.204 |
|  | PP>PPc | 0.586 | 2.016 | 1.796 | -3.366 | 4.537 | 0.290 | 0.084 | 0.771 |
| R IFG | (Intercept) | -0.378 | 0.37 | 0.686 | -1.102 | 0.347 | -1.022 | 1.044 | 0.307 |
|  | AP>Apc | 1.310 | 1.110 | 3.705 | -0.865 | 3.484 | 1.180 | 1.393 | 0.238 |
|  | PP>PPc | 3.772 | 1.702 | 43.464 | 0.436 | 7.108 | 2.216 | 4.911 | 0.027* |
| R SPL | (Intercept) | -0.112 | 0.334 | 0.894 | -0.767 | 0.543 | -0.336 | 0.113 | 0.737 |
|  | AP>APc | -0.463 | 0.983 | 0.629 | -2.39 | 1.464 | -0.471 | 0.222 | 0.638 |
|  | PP>PPc | 1.949 | 1.205 | 7.024 | -0.411 | 4.31 | 1.618 | 2.619 | 0.106 |
| R SM | (Intercept) | -0.190 | 0.383 | 0.827 | -0.941 | 0.561 | -0.496 | 0.246 | 0.620 |
|  | AP>APc | -1.640 | 1.354 | 0.194 | -4.294 | 1.013 | -1.211 | 1.468 | 0.226 |
|  | PP>PPc | 1.460 | 1.686 | 4.305 | -1.845 | 4.765 | 0.866 | 0.749 | 0.387 |

The linear regression models employing the beta values of the contrasts of interest (AP > APc and PP > PPc) extracted from R and L IFG as dependent variables and BDI, PCS, and BES scores (the psychometric measures shown to be different between the two groups) as predictors, were not statistically significant (see Table 6).

**Table 6.** Linear regression models results. For each significant ROI from the logistic regression model (i.e, right and left IFG) and for each contrast (AP > APc, PP > PP) a linear regression model was built to evaluate the association between the extracted beta values and the predictors of interest (PCS, BDI, BES). Abbreviations: PCS, Pain Catastrophizing Scale; BDI, Beck Depression Inventory; AP, affective empathy condition; APc, control condition for affective empathy; PP, physical empathy condition; PPc, control for physical empathy.

|  | R IFG |  |  | L IFG |  |  |
| --- | --- | --- | --- | --- | --- | --- |
| Model summary |  |  |  |  |  |  |
|  | R<sup>2</sup> | F (df) | p-value | R<sup>2</sup> | F(df) | p-value |
| AP>APc | 0.033 | 0.416(3) | 0.743 | 0.067 | 0.883(3) | 0.459 |
| PP>PPc | 0.077 | 1.031(3) | 0.390 | 0.028 | 0.354(3) | 0.787 |
| Coefficients |  |  |  |  |  |  |
| Predictors | Beta | t | p | Beta | t | p |
| AP>APc |  |  |  |  |  |  |
| (Intercept) |  | -0.480 | 0.634 |  | -0.331 | 0.742 |
| PCS | -0.045 | -0.234 | 0.817 | -0.202 | -1.073 | 0.290 |
| BDI | 0.188 | 0.945 | 0.351 | 0.305 | 1.562 | 0.127 |
| BES | 0.030 | 0.177 | 0.861 | -0.012 | -0.071 | 0.944 |
| PP>PPc |  |  |  |  |  |  |
| (Intercept) |  | 0.854 | 0.040 |  | 0.544 | 0.590 |
| PCS | 0.265 | 1.419 | 0.164 | 0.115 | 0.602 | 0.551 |
| BDI | -0.007 | -0.036 | 0.971 | -0.196 | -0.984 | 0.332 |
| BES | -0.158 | -0.934 | 0.352 | -0.002 | -0.014 | 0.989 |

The correlational analyses showed that during physical pain empathy (contrast PP vs PPc), the beta values extracted from the R IFG were significantly correlated with the level of pain experienced during the attacks (r =.458, p = 0.037) (see Table 7). No other significant correlation was observed.

**Table 7.** Correlational analyses between the significant ROIs beta values (obtained from the logistic regression models) and clinical variables. * Significant p-value: p< 0.05. Abbreviations: IFG, inferior frontal gyrus; L, left; R, right; AP, affective empathy condition; APc, control condition for affective empathy; PP, physical empathy condition; PPc, control for physical empathy.

| *Variable* | **R IFG** |  |  |  | **L IFG** |  |  |  |
| --- | --- | --- | --- | --- | --- | --- | --- | --- |
|  | **AP>AC** |  | **PP>PC** |  | **AP>AC** |  | **PP>PC** |  |
|  | *Statistic* | *p-value* | *Statistic* | *p-value* | *Statistic* | *p-value* | *Statistic* | *p-value* |
| Disease duration | r=-0.018 | 0.937 | r=0.057 | 0.807 | r=0.139 | 0.548 | r=-0.31 | 0.171 |
| Pain level during attacks<br>(VAS: 0-10) | r=-0.300 | 0.186 | r=0.458 | 0.037* | r=-0.253 | 0.269 | r=0.216 | 0.346 |

### 3.3. Vicarious Pain Signature (VPS): predicting affective (AP) and physical pain (PP) empathy in MIG and CTRL group

We found that the FE-VPS (i.e., facial expression vicarious pain) and NS-VPS (i.e., noxious stimulation vicarious pain) reached high prediction accuracy in both groups in distinguishing empathy for pain (i.e., AP or PP) with respect to the respective control condition (i.e., APc or PPc).

As shown in Figure 4 and Table 8, NS-VPS (i.e., noxious stimulation vicarious pain) was able to differentiate PP and PPc with high accuracy in both MIG patients [n = 42, accuracy 100±0.0% SE, Cohen’s d = 2.59, sensitivity 100%, specificity: 100%, two-sided binomial test: p < 0.001] and CTRL group [n = 42, accuracy 86%±7.6% (SE), Cohen’s d = 1.23, sensitivity 86%, specificity: 86%, two-sided binomial test: p = 0.001].

**Figure 4:**
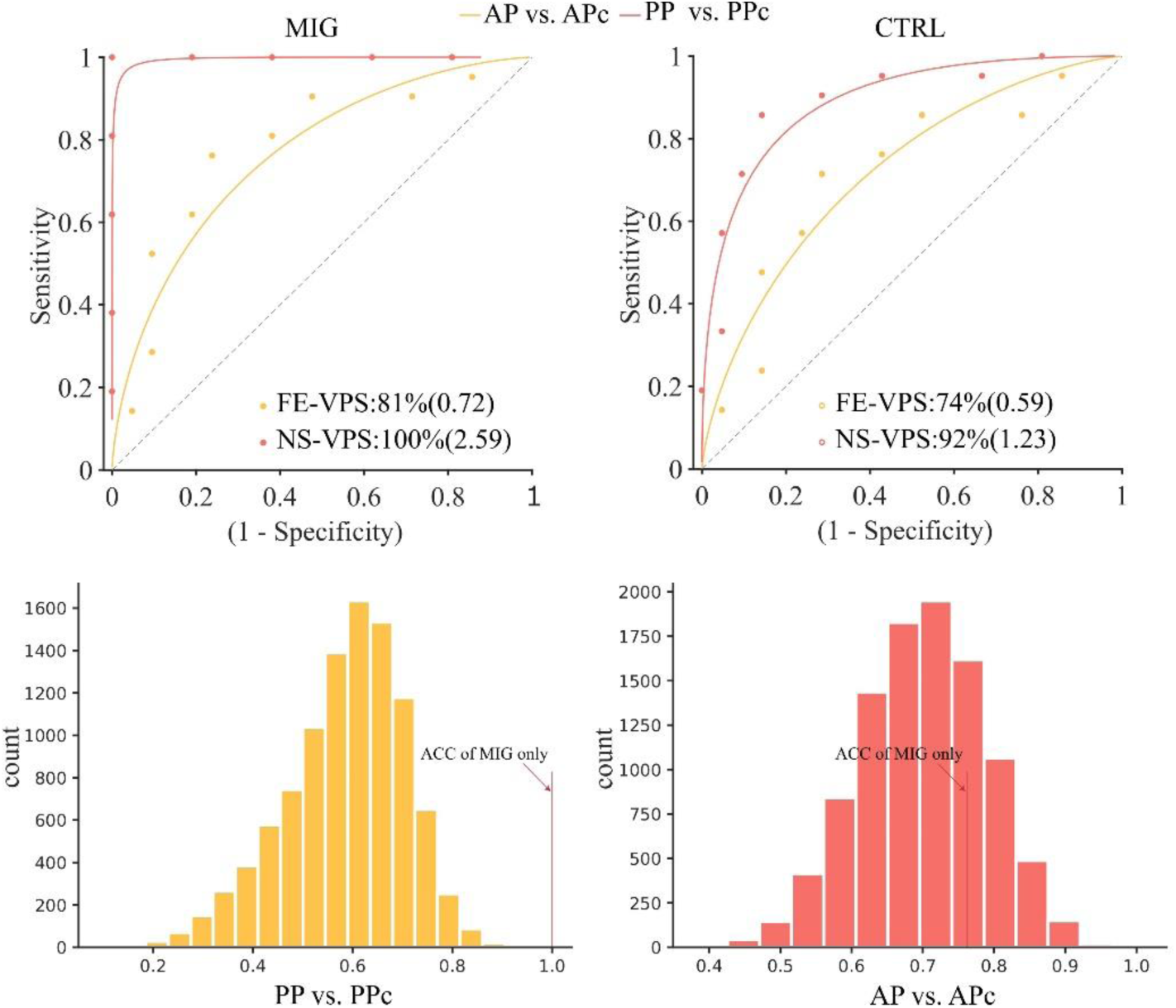
A. Evaluation of VPS (Vicarious Pain Signature) (Zhou et al., 2020) predictive patterns using ROC analysis in migraine and control groups for affective pain empathy (employing the VPS for facial expression vicarious pain or FE) and physical pain empathy (employing the VPS for noxious stimulation vicarious pain or NS); B. Null distribution of classification accuracy values (10000 permutations). The observed accuracy was compared to this null distribution to calculate empirical p-values. Abbreviations: ACC, accuracy; PP, physical pain empathy; PPc, physical pain empathy control; AP, affective pain empathy; APc, affective pain empathy control.

**Table 8.** Performance metrics for the classification of affective pain (AP vs. APc) and physical pain (PP vs. PPc) conditions in the MIG and CTRL groups using the facial expression vicarious pain signature (FE-VPS) and noxious stimulation vicarious pain (NS-VPS) developed by Zhou et al. (2020). Abbreviations: sens, sensitivity; spec, specificity; PPC, positive predictive value; CI, 95% confidence intervals. Nonparametric and parametric AUC values indicate the area under the curve as a measure of classifier accuracy.

| **Comparison** |  | **Sens (95% CI)** | **Spec (95% CI)** | **PPV (95% CI)** | **AUC** | **Cohen's d** | **Accuracy ± SE** | **p-value** |
| --- | --- | --- | --- | --- | --- | --- | --- | --- |
| **MIG** | **FE-VPS (AP vs. APc)** | 76% (56–94%) | 76% (57–94%) | 76% (57–94%) | 0.81 | 0.72 | 76% ± 9.3% | 0.027 |
| | **NS-VPS (PP vs. PPc)** | 100% (100–100%) | 100% (100–100%) | 100% (100–100%) | 1.00 | 2.59 | 100% ± 0.0% | 0.000 |
| **CTRL** | **FE-VPS (AP vs. APc)** | 71% (50–90%) | 71% (52–89%) | 71% (52–89%) | 0.74 | 0.59 | 71% ± 9.9% | 0.078 |
| | **NS-VPS (PP vs. PC)** | 86% (70–100%) | 86% (69–100%) | 86% (68–100%) | 0.92 | 1.23 | 86% ± 7.6% | 0.001 |

FE-VPS (i.e., facial expression vicarious pain) could reach high accuracy between AP vs. APc in MIG patients [accuracy 76%±9.3% SE, Cohen’s d = 0.72, sensitivity: 76%, specificity 76%, two-sided binomial test P = 0.026] with a trend towards significance for CTRL group [accuracy: 71%±9.9% (SE), Cohen’s d =0.59, sensitivity 71%, specificity: 71%, two-sided binomial test: p = 0.078]. Further permutation test confirmed that the observed accuracy for MIG patients for the physical empathy for pain (p*=*0.036), but not for the affective empathy for pain (p*=*0.711), was significantly higher than expected under the null hypothesis (see Figure 4), thus robustly indicating that there are significant differences between the pattern expression of the two groups.

## 4. Discussion

Using a well-validated pain empathic fMRI paradigm (J. Li et al. 2019; X. Xu et al. 2020; Zhou et al. 2020), we studied the activity of key brain regions of empathy for pain processing during physical and affective stimuli in a group of MIG patients, comparing them with those in a control group. Through the application of robust statistical methods, including logistic regression analyses, an unbiased approach to select the ROIs and whole brain multivariate pattern expression, our results revealed that MIG patients exhibit increased neural reactivity in the bilateral IFG during empathy for physical pain with stronger neurofunctional alterations in the right IFG being related to higher levels of pain experienced during the migraine attacks. Further strengthening these results, the whole-brain predictive model for vicarious pain for noxious stimulation (NS-VPS) developed by Zhou et al. (2020) applied to the fMRI maps (i.e., con images) of MIG patients achieved an accuracy of 100% in correctly classifying physical pain empathy in respect to the control conditions, while a more limited but still significant accuracy (86%) was achieved for the CTRL group. Remarkably, the predictive accuracy for MIG patients for physical pain empathy, but not for affective pain empathy, has been demonstrated to be significantly different from the pattern expression of the MIG and CTRL groups together, indicating a more robust neural representation of physical pain empathy in the MIG group in respect to the CTRL group on the whole brain level. In contrast, neither regional nor subtle whole brain alterations were observed during affective pain empathic processing, indicating domain-specific alterations.

Altogether, these results robustly indicate a heightened and domain-specific neural sensitivity to physical pain empathy in patients with migraine compared to the CTRL group, in particular in the bilateral IFG. The IFG is a broad and not uniform region anatomically, cytoarchitectonically, and functionally (Amunts et al. 1999; Wojtasik et al. 2020). It is involved in a plethora of processes: while the left IFG is known to be involved in semantic and phonological processing (Liakakis, Nickel, and Seitz 2011), action observation, and imitation (Hamzei et al. 2016), the right IFG is thought to be a cardinal region of social cognition, emotional and interoceptive processes (Adolfi et al. 2017), body awareness (Luo et al. 2022), and regulatory processes (Zhuang et al. 2023). More recently, IFG has been shown to be causally involved in the determination of the conscious experience (Weilnhammer et al. 2021).

In the context of interest for this investigation, a recent meta-analysis of fMRI studies identified the IFG as a crucial node in the empathy-for-pain network (Fallon, Roberts, and Stancak 2020). Delving deeper into the role of this region in empathy for pain, it was recently shown that the right IFG is causally involved in empathy for physical pain, suggesting a role in the encoding of pain through visual cues that would allow inference of the other’s state (Y. Li et al. 2021). Furthermore, a series of real-time fMRI neurofeedback investigations employing the reappraisal technique highlighted the regulatory role of this region for physical pain empathy (Naor et al. 2020) in combination with the anterior insula (Yao et al. 2016; Y. Zhang et al. 2023).

As discussed in the introductory section, we now have multiple pieces of evidence demonstrating a good overlap of the neural pathways involved in empathy for pain and nociceptive processing. In fact, in agreement with previous works (Singer et al. 2004; Lamm, Decety, and Singer 2011; Timmers et al. 2018), the meta-analysis of Fallon et al. (2020) demonstrated the presence of shared neural pathways of these two processes and showed that the bilateral IFG is one of the hub regions involved in both processes. This commonality of neural pathways was also elegantly demonstrated by studies employing multivoxel pattern analyses, which showed that the neural patterns capable of predicting the processing of stimuli that elicited empathy for physical pain were also capable of predicting the processing of acute pain and vice versa (Zhou et al. 2020; M. Li et al. 2024).

Based on the above studies, two inferences can be made in the context of the heightened responsivity observed in MIG patients in the bilateral IFG during empathy for physical pain. First, an abnormal IFG activation could reflect a regulatory strategy in the face of increased allostatic load, particularly during empathy for physical pain. In this case, the IFG might be involved in modulating the response to these aversive stimuli, suggesting that patients with MIG might exhibit a more intense response to such stimulation. However, it is important to note that as no alterations were found in empathy for emotional pain, this hypothesis is relatively weak, although an increased salience of images showing pain evoked by physical stimuli cannot be excluded. Second, from a translational perspective, the increased reactivity of the IFG would seem to be strongly indicative of a specific alteration of the pathways involved in the pain response. This could indicate that the IFG regulates a system that is hyperresponsive to physical pain signals, as suggested by the specific activation during empathy for pain, which emulates neural patterns similar to those observed during nociception (Zhou et al. 2020; M. Li et al. 2024). This hypothesis is also supported by the significant correlation observed between the level of pain experienced during the attacks and the activity observed in the R IFG during physical pain empathy.

In agreement with the last hypothesis, long-term pain experiences are thought to induce maladaptive changes in neural processes associated with empathy for pain; however, relatively few studies have investigated the neural basis of this process in pain-related disorders showing results of different signs.

In primary dysmenorrhea, two complementary studies (Mu et al. 2021; C. Wang et al. 2024) revealed heightened behavioral sensitivity to pain empathy alongside structural changes in the insular cortex and somatosensory areas, which were identified as mediators between menstrual pain intensity and pain empathy for pain. In these studies, empathy for pain was triggered by the observation of noxious stimulation of body limbs, thus with stimuli triggering the physical dimension of empathy for pain. In contrast, fibromyalgia, a condition primarily characterized by widespread musculoskeletal pain and heightened sensitivity to pain (Burgmer et al. 2009; Müller et al. 2021) has been associated with decreased activity in several brain regions during empathy for physical pain (S. J. Lee et al. 2013).

It is important to note that empathy for pain is considered a disorder-specific as well as also a potentially transdiagnostic biomarker of neuropsychiatric disorders, including depression (X. Xu et al. 2020; He et al. 2024) with which migraine has a high comorbidity. Our results contrast sharply with the reduced brain responses to empathy for physical pain observed in a major depressive disorder study, which employed the same experimental fMRI paradigm as ours (X. Xu et al. 2020). Consistent with the previous literature (Caponnetto et al. 2021; Irimia et al. 2021), the MIG patients in our sample exhibited increased levels of depressive symptoms; however, the above apparent dissociation suggests a specific brain activity in MIG patients that can distinguish them from depressed subjects, thus indicating the need for more targeted investigations into the neurobiological mechanisms underlying the comorbidity between migraine and depression.

From a clinical perspective, the MIG patients presented a robust increase in pain catastrophizing scores (i.e. at the PCS scale). Pain catastrophizing is characterized by an overmagnification of the threat associated with pain, a sense of helplessness, and a relative inability to prevent or inhibit pain-related thoughts, even outside the painful event. In other words, it refers to a person’s tendency to anticipate and perceive pain in an exaggerated way and respond with intense emotions and negative thoughts (Quartana, Campbell, and Edwards 2009). In our sample, patients with MIG showed, on average, pain catastrophizing scores (M±SD: 35.0 ± 11.98) above the cuto*ff* for clinical relevance (Sullivan, Bishop, and Pivik 1995) with a particular exacerbation in the sub-scales of impotence and rumination. This strong attitude to pain catastrophizing partly contrasts with previous studies reporting increased PCS scores in MIG patients but without reaching clinical relevance (Kim et al. 2021; Galioto et al. 2017). Our results are more in line with those reported in chronic MIG patients with medication overuse (Grazzi et al. 2023) and in MIG patients with obesity (Bond et al. 2015), who showed clinically relevant pain catastrophization. Interestingly, our results also showed that patients with MIG presented slightly higher scores on the empathy scale (i.e., BES) than the CTRL group, with a significant difference in the cognitive dimension of empathy. Although not central to this work, it is important to note that excessive empathy for others confers a vulnerability to emotional disorders possibly due to the excessive allostatic load (Chikovani et al. 2015).

Despite the interesting results, it is important to note that our work has some limitations. The most important one is the relatively limited sample of MIG patients. However, the abnormal responses specific to physical pain empathy were robustly supported by the application of the VPS-NP (i.e., Vicarious Pain Signature for nociceptive pain), which achieved 100% accuracy in predicting the empathy for physical pain stimulation with respect to the control condition. A second important limitation is that we excluded only patients who presented an attack 24 hours before the fMRI scans. However, in this case, it is important to note that the MIG patients of our sample presented a relatively low number of attacks per month in the range 1-5, thus limiting the possibility that they were not in the interictal phase. As the last limitation, we have to mention that it is possible that physical pain empathy and affective pain empathy stimuli might present different saliency, thus the dissociation between the two dimensions of empathy for pain (i.e., physical and affective) can be an effect of the different allostatic load of the two stimuli.

Contrary to our hypothesis of similar abnormal neural responses across the two investigated dimensions of empathy for pain (physical and emotional), our results indicate that MIG patients present an increased brain reactivity, mainly involving the bilateral IFG, during empathy for physical pain stimuli. Based on the fact that the neural pathways involved in empathy for physical pain and in acute pain processing robustly overlap, these results point to a specific hypersensitization of the pain pathways. Moreover, our results do not suggest a general hyperresponsive brain to aversive stimuli, but rather an increased brain responsiveness specifically related to pain processing. The apparent dissociation of the neural pattern evidenced in a study on major depressive disorder (X. Xu et al. 2020) and that employed the same experimental paradigm as in our study calls for a deeper analysis of the relationship between migraine and depression.

## Author Approval

All authors have seen and approved the manuscript.

## Data availability

Data will be made available upon reasonable request to the corresponding author.

## Data Availability

All data produced in the present study are available upon reasonable request to the authors

