## Supplemental Materials for "Regional and whole-brain neurofunctional alterations during pain empathic processing of physical but not affective pain in migraine patients"

| Hemisphere | Brain Region | K_E_ | pFWE | F | Z_E_ | Coordinate (MNI) |
| --- | --- | --- | --- | --- | --- | --- |
| L | Temp Inf, Temp Mid, Occipital Mid, Occipital Sup, Parietal Sup | 4014 | 0.0000 | 94.19 | Inf | [-51;-64;-7] |
| R | Temp Inf, Temp Mid, Occipital Mid, Occipital Sup, Parietal Sup | 2827 | 0.0000 | 61.14 | Inf | [36;-79;17] |
| L | Front Sup, Front Mid, Precentral | 201 | 0.0000 | 40.53 | Inf | [-24;-7;53] |
| L | Precentral, Front Inf Oper, Rolandic Oper | 273 | 0.0000 | 35.66 | Inf | [-54;5;32] |
| L | Angular | 50 | 0.0000 | 22.89 | 6.97 | [-48;-73;41] |
| L | Front Inf Tri | 67 | 0.0001 | 16.53 | 5.91 | [-48;32;14] |
| R | Supramarginal, Parietal Inf | 45 | 0.0001 | 16.08 | 5.82 | [30;-43;-40] |
| R | Precentral | 27 | 0.0006 | 14.55 | 5.52 | [30;-4;53] |
| R | Cerebellum | 12 | 0.0017 | 13.53 | 5.30 | [12;-76;-40] |
| L | Thalamus | 14 | 0.0086 | 12.00 | 4.96 | [-15;-25;8] |

**Table 1 SM.** Results of the whole-brain analysis (MNI coordinates of the significant clusters). The main effect of the condition factor was obtained from a full factorial design with ‘condition’ (AP, APc, PP, PPc) as the within-subject factor and ‘group’ (MIG, CTRL) as the between-subject factor, using FWE-corrected p-values at 0.05.

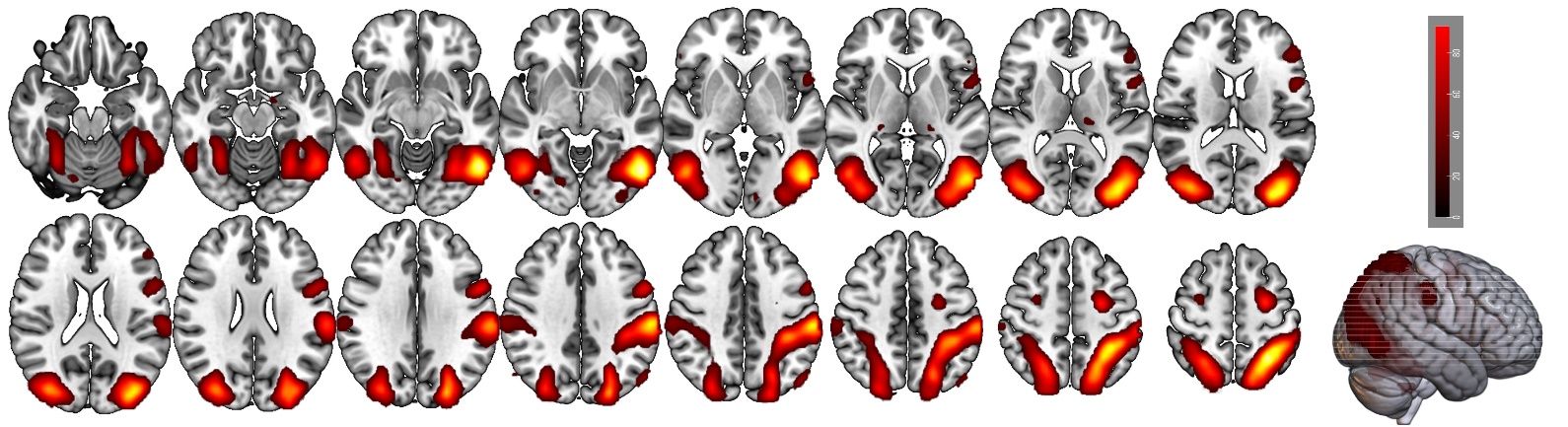

**Figure 1 SM**: Results of the whole-brain analysis. The main effect of the condition factor was obtained from a full factorial design with ‘condition’ (AP, APc, PP, PPc) as the within-subject factor and ‘group’ (MIG, CTRL) as the between-subject factor, using FWE-corrected p-values at 0.05. The color scale represents the F-values.
